# Sexual Identity Moderates Age, Cigarette Use, and Depressive Symptom Trajectories Across Developmental Age

**DOI:** 10.1101/2025.06.18.25329725

**Authors:** Taylor R. Harrington, Gia E. Barboza-Salerno, Joanne G. Patterson

**Affiliations:** The Ohio State University College of Public Health, 1841 Ave. Columbus, OH, 43210; The Ohio State University College of Social Work, 1947 College Rd. N Columbus, OH, 43210

**Keywords:** adolescents, depression, trajectory, cigarette use, SGM

## Abstract

**Background:** Evidence linking smoking and depressive symptoms is inconsistent, warranting longitudinal studies using robust methods to assess causal relationships.

**Study Design:** Group-based trajectory models were used to investigate associations between cigarette use and depressive symptoms across developmental age and the moderating effects of sexual identity.

**Methods:** Multilevel modeling was used to compare trajectories of depressive symptoms from the Adolescent Health Survey. Regions of significance were estimated using Johnson-Neyman significance intervals for interaction terms. We modeled depressive symptom trajectories as a function of age, smoking behavior, and sexual orientation interaction.

**Results:** The conditional model yielded an ICC of 0.34, indicating a strong clustering effect over time. Both age (β□= 0.069, p□<□0.001) and age-squared (β□= -0.002, p□<□0.001) were positively associated with initial values of depressive symptoms, demonstrating significantly steeper declines over time (βage = -0.001, p□<□0.001). Bisexual identification was positively associated with depressive symptoms (β□= 0.001, p□<□0.01) and cigarette use (β□= 0.014, p□<□0.001) trajectories were significantly higher among bisexual identities up through young adulthood.

**Conclusions:** Our result highlights the importance of focusing interventions for those of bisexual identity in adolescence and their long-term impact on bisexual populations who use cigarettes.

- Bisexual individuals experience distinct differences regarding smoking behaviors and depressive symptoms across developmental age.
- It is essential to stratify identities into their respective groups rather than conflating them into a sexual minority group identity.
- Early-life interventions are necessary to support minoritized populations, particularly those who identify as bisexual.
- Specific implementation of policies to improve mental health and smoking cessation for those identifying as bisexual is highly suggested.

## Introduction

In the US, an estimated 8% of adults have clinical depression, and 11.3% experience depression weekly.^1^ Between 2005 and 2006, 5.4% of Americans over the age of 12 experienced depression.^2^ The observed increased levels of depressive symptoms among adolescents (between the ages of 10 and 17) and young adults (ages of 18 to 25) are of public health concern.^3,4^ Depression is known to be associated with an increased likelihood of smoking tobacco, illicit drug use/abuse, alcohol abuse, lower levels of physical activity and fitness, higher levels of obesity, higher risk of cardiovascular disease, and lower levels of quality of life.^1^ Although an association of depression with tobacco use is noted in the literature,^1,5–10^ no research to date has examined the association between smoking and depressive symptom trajectories across developmental age. Accordingly, we utilize growth mixture modeling to identify this relationship and investigate how sexual identity may moderate the relationship.

### Depressive Symptom Trajectories across Developmental Age

Adolescents and young adults disproportionately experience more depressive symptoms than their middle-aged counterparts. The global burden of disease of mental disorders for those between the ages of 10 and 19 is 13%. Additionally, mental health is still largely underrecognized and untreated.^11^ Depressive symptoms follow a U-shaped curve throughout the life course; they are high in adolescence, decrease with the transition to adulthood, and increase again in older adults.^12,13^ This trend is linked to the stressors experienced during these life stages; adolescence in particular is filled with challenging transformations across physical, emotional, developmental, and social domains.^12^

Although depressive symptoms are dynamic throughout the life course, disparities are prevalent in early adolescence and only increase with age into adulthood.^12^ In young adults, there is substantial heterogeneity in the life course of depressive symptoms.^10^ The use of trajectory models aids in deciphering this heterogeneity.^10^ Obtaining knowledge of developmental timing would provide relevant information for the planning and timing of interventions.^14^ Lastly, identification of trajectory-specific risk and protective factors could shed light on the etiological underpinnings of depressive symptom courses.^14^ Known predictors of elevated depressive symptom trajectories include female gender, ethnic minority status, delinquent behavior, and substance use in adolescence.^14^ This study adds to the literature by investigating the specific risk factor of cigarette use as a predictor of depressive symptom trajectories across developmental age.

### Associations Between Cigarette Use and Depressive Symptoms

Individuals with mental health challenges disproportionately experience higher rates of smoking. From 2012 to 2014, smoking prevalence for those with a past-year mental illness was 33%, compared to 21% for those without ^9,15^ In the past few decades, the trend of smoking has declined, yet this has not been observed for individuals with mental illnesses.^9^ Friedman and colleagues found that individuals with poorer mental health have higher odds of initiating first-time cigarette use.^9^ For current smokers, there are increased odds of having mental health symptoms even when controlling for age and prior mental health treatment, consistent across gender.^7^ Additionally, regular tobacco use (smoking before age 13) is found to be significantly and positively associated with a lifetime diagnosis of major depressive disorder.^7^ Investigation of adolescent and young adult depressive symptoms trajectories and cigarette use is imperative.

### Sexual Identity as a Moderator of the Association Between Cigarette Use and Depressive Symptom Trajectories

Cigarette use is predominant across all sexual and gender minorities (SGM) which includes members of lesbian, gay, bisexual, transgender, and queer (LGBTQ) communities.^16,17^ In the United States, between 30 and 38 percent of SGM adults reported current cigarette use compared to 16 to 20 percent of heterosexual adults.^2^ SGM tobacco users are reported to be less successful in quitting compared to their heterosexual counterparts, furthering the disparity already experienced by SGM.^2^

Sexual minorities (SM) report poorer mental health, including higher rates of depression and depressive symptoms, compared to non-sexual minorities.^18^ In fact, a systematic review found a consistent pattern of low rates of depression among heterosexual individuals, where bisexual individuals demonstrate greater or equivalent rates to lesbian or gay individuals.^19^ *Linking Smoking Behavior to Depressive Symptoms: Theoretical Framework*

According to the minority stress theory (MST), SGM health disparities exist from excess exposure to social stress, arising from sexual orientation-based stigma, contributing to tobacco use disparities.^16,20,21^ MST is characterized by stigmas such as discriminatory policies, major life events, interpersonal discrimination, victimization, and microaggressions.^21^ These stressors create an excess burden on SGMs, putting them at greater risk of negative health outcomes compared to their counterparts.^21^ Stigma-related stress can lead to an increase in emotional dysregulation, social/interpersonal issues, and risk for psychopathology.^21^ Young adults who are of SGM are at the highest risk of tobacco initiation in young adulthood.^20^ Thus, SGM individuals who experience minority related stress demonstrate poorer mental health outcomes than those who do not face similar levels of stress.^22^

### Current Study

No prior research has directly examined the impact of cigarette use as a predictor of depressive symptoms in adolescence, and whether these trajectories vary for SGM (vs heterosexual) youth. Gaining an understanding of this relationship is critical to reducing disparities in mental health, cigarette use, and to determining the best windows of opportunity across the lifespan for interventions. Consequently, the purpose of this study is to assess how cigarette use affects the trajectories of depression, specifically investigating how sexual identity moderates this relationship.

Longitudinal studies of cigarette smoking have demonstrated a bell curve pattern, of which there is an increase in use during adolescence, followed by stability to the late teens and onwards throughout the life course. More recently, researchers are utilizing the person- and variable-centered approaches in longitudinal studies.^8^ In doing so, there is the ability to examine multiple developmental pathways that have previously only been observed through individual heterogeneity in smoking behaviors.^8^ This strategy organizes the individual characteristics in a meaningful way that highlights subgroups of people. Through the process of grouping factors into different depression trajectories, the combined person- and variable-centered analysis may inform prevention programs that are best tailored for subgroups within a population.^8^ Accordingly, we investigated depressive symptoms utilizing a person-centered approach. We analyzed data drawn from the large, nationally representative survey, National Longitudinal Study of Adolescent Health (Add Health), and applied trajectory modeling to 1. Determine how cigarette use may predict depression symptoms across the life course, and 2. Determine how sexual identity moderates the relationship.

## Methods

### Participants

The data for the study and analyses were drawn from Add Health; a nationally representative, probability-based survey that examined a broad range of health-related attitudes and behaviors of U.S. adolescents, which can be found here (https://addhealth.cpc.unc.edu/data/#public-use).^8^ A systematic random sample of 80 high schools was selected proportional to enrollment size and stratified by geographic region, urbanicity, school type, and ethnic mix.^8^ For this secondary analysis, we included four waves of data: baseline, 1-year post-baseline, 5 years post-baseline, and 14 years post-baseline. The final age of the sample participants ranged from 24 to 32 years.^15^ The mean ages of the participants at each of the four waves of data collection were 15.65 (SD = 1.75), 16.22 (SD = 1.64), 22.96 (SD = 1.77), and 28.9 (SD = 1.76).

The Add Health study captured data from adolescents in multiple overlapping age cohorts, which allows for the examination of developmental trajectories spanning early adolescence to young adulthood. This is accomplished by “linking” the cohorts together based on age in a cohort-sequential design. Group-based trajectory models cannot accommodate cases with missing covariates; thus, sixty-four participants were missing smoking outcome measures at all assessment points and were not included in our analyses. We conducted our analyses with the remaining 10,720 participants who completed an in-home survey.

### Procedure

For the in-home survey, an interviewer read the questions aloud and recorded the respondent’s answers on a laptop using a computer-assisted personal interview system. Parts of the survey with potentially sensitive information were administered using an audio computer-assisted self-interview that allowed the participants to enter their responses directly into the computer. For this analysis, we obtained secondary data, and the four waves were linked by participant age.

### Measures

#### Demographic characteristics

##### Age

Adolescents’ age in years was calculated for Wave 1 by using their birthdate at the time of the first interview date. The age is computed by finding the difference between the interview date and the birth date. Waves 2 and 3 provided calculated age variables. For Wave 4, we carried out the same process as for Wave 1.

##### Gender

Wave 1 measured biological sex (1= males, 1=females). The final wave (wave 4) included questions regarding sexual orientation and gender identity. Our sample consisted of 56.27% females and 43.73% males.

#### Moderating Factors

##### Cigarette Use

At each Wave, participants were asked to report the number of cigarettes (quantity) and frequency (number of days) of smoking in the past 30 days to capture the diversity of smoking patterns in this population. For example, to differentiate between a participant who smokes only one cigarette per day for 30 days versus a participant who smokes 30 cigarettes in one day and not again for the rest of that month. Following Costello^5^ and colleagues’ approach, we categorized the variables as followed: 1) quantity: 0, 1-5, 6-10, 11-20, 20+; and 2) frequency: 0, 1-5, 6-10, 11-15, 16-20, 21-25, 26-29, 30 (5). The categorical quantity and frequency variables’ values were then multiplied to obtain a single value, ranging from 0 to 28. Creating the variable in this way accurately captures diverse patterns of smoking behavior. The mean value across all four waves was 6.83 (SD = 6.44).

##### Sexual Identity

Sexual identity was asked in the last two survey waves (waves 3 and 4) as follows: “Please choose the description that best fits how you think about yourself:

1. 100% heterosexual (straight)
2. Mostly heterosexual (straight) but somewhat attracted to people of your own sex
3. Bisexual, that is, attracted to men and women equally
4. Mostly homosexual (gay), but somewhat attracted to people of the opposite sex
5. 100% homosexual (gay)

Following Sabia’s ^23^ coding of sexual orientation, individuals who responded being “100% heterosexual” were coded as straight. Those who chose “Mostly heterosexual”, “Bisexual”, and “Mostly Homosexual” were coded as bisexual. Those who reported being “100% homosexual” were coded as gay/lesbian.

#### Outcome Measure

##### Depression

Measured at Wave 1, 20 items correspond to the Center for Epidemiological Studies – Depression Scale (CES-D). Responses are presented using a 4-point scale (0=never or rarely and 3=most of the time or all of the time) to indicate how often participants felt emotional states (“felt depressed”, “felt happy”) in the past week. Items are then summed to develop a total score for each adolescent. The CES-D scores range from 0 to 60, with higher scores demonstrating the presence of more symptomatology.^24^ The CES-D used in the Add Health survey has been shortened; however, it has demonstrated previous adequate reliability (Cronbach’s α =0.79) and validity. The values of depression from all four waves ranged from 0 to 21, and the mean score was 5.72 (SD = 3.06) (Cronbach’s α=0.61).

### Analytic Strategy

We used a growth curve mixture model method to identify depression trajectories across developmental ages (ranging from 13 to 35 years old). With the utilization of the “lmer” function from the “lme4” package in R, we fit a linear mixed-effects model where depression scores were the dependent variable and standardized to have a mean of zero and a standard deviation of one. Multilevel modeling was used to compare trajectories of depressive symptoms by structuring four waves of data from the Adolescent Health Survey to reflect developmental age (12-31 years). The *lmer* function in R was used to test multiple linear mixed models using restricted maximum likelihood estimation. Regions of significance were estimated using Johnson-Neyman significance intervals for interaction terms. The best fitting model, determined by the Akaike Information Criterion, described depressive symptom trajectories as a function of age, age-squared, cigarette use, sexual identity, and cigarette use by sexual identity interaction.

## Results

We began with the intercept-only model (“Model 0” in Table 2), which included depressive scores as the standardized dependent variable that has a mean of 0 and a standard deviation of 1. One predictor variable is age, which is included in the fixed effects of the model. Another predictor is the quadratic effect of age (age^2^). Lastly, we applied a random intercept for participants’ ID, allowing each participant to have their own intercept to account for the variability between participants. Model 0 had the largest BIC (29001.70), suggesting there is another model that better fits the data. Adding smoking behavior as a predictor (“Model 1” in Table 2) to Model 0 decreased the BIC (28757.52), signifying that the inclusion of smoking as a predictor of depressive symptoms improved the model fit. Thus, smoking behavior is an important, statistically significant (p < 0.001) predictor of depressive symptoms. In Model 2, the interaction between age and sexual identity was added to assess how the effect of age on depressive symptoms differs by sexual identity. Model 2 produced the lowest BIC (26453.923) of all the models, suggesting the effect of age on depressive symptoms varies depending on one’s sexual identity. Essentially, the relationship between age and depressive symptoms is not uniform across sexual identity. Identification as straight was statistically significant (p < 0.001), while identification as gay/lesbian (p = 0.114) was not. Lastly, in Model 3, a three-way interaction between age, smoking behavior, and sexual identity was added, allowing us to assess the effect of age on depressive symptoms and allow it to vary depending on both sexual identity and smoking behavior. Model 3 provided a low BIC (26479.748); however, it was slightly higher than the BIC from Model 2. Thus, still conclude Model 2 as the best fitting model for the data. Noteworthy, in Model 3, bisexual sexual identity was significant (p = 0.003), suggesting that individuals who identify as bisexual experience a different relationship between age and depressive symptoms compared to individuals with other sexual identities. This also suggests that the effect is conditional on one’s smoking behavior. All models yielded an ICC of 0.34, demonstrating a strong clustering effect over time.

**Table 1:** Participant Demographic Characteristics by Sexual Orientation.

| Demographics | **Bisexual, N = 304<sup>1</sup>** | **Gay/Lesbian, N = 52<sup>1</sup>** | **Straight, N = 9,604** | **Missing, N = 760** |
| --- | --- | --- | --- | --- |
| Age | 19.0 (16.0, 26.0) | 19.0 (15.8, 24.5) | 19.0 (16.0, 26.0) | 19.0 (16.0, 25.0) |
| Sex |  |  |  |  |
| Female | 234 (77%) | 12 (23%) | 5,228 (54%) | 558 (73%) |
| Male | 70 (23%) | 40 (77%) | 4,376 (46%) | 202 (27%) |
| Race |  |  |  |  |
| White non-Hispanic | 222 (73%) | 44 (85%) | 6,680 (70%) | 558 (73%) |
| African American | 49 (16%) | 4 (7.7%) | 2,020 (21%) | 127 (17%) |
| Asian non-Hispanic | 9 (3.0%) | 0 (0%) | 260 (2.7%) | 15 (2.0%) |
| Hispanic all races | 0 (0%) | 0 (0%) | 16 (0.2%) | 4 (0.5%) |
| Native American | 10 (3.3%) | 0 (0%) | 160 (1.7%) | 14 (1.8%) |
| Other non-Hispanic | 14 (4.6%) | 4 (7.7%) | 468 (4.9%) | 42 (5.5%) |
| Depressive Symptoms | 6.0 (4.0, 8.0) | 5.5 (4.0, 8.0) | 5.0 (3.0, 7.0) | 6.0 (4.0, 9.0) |
| Smoking | 4 (4, 10) | 4 (4, 7) | 4 (4, 4) | 4 (4, 8) |

<sup>1</sup>Median (IQR); %

**Table 2:** Overview of Linear Mixed-Effects Models with Interaction Effects on Depression.

|  | Model 0 | Model 1 | Model 2 | **Model 3** |
| --- | --- | --- | --- | --- |
| (Intercept) | -0.729 *** | -0.638 *** | -0.624 *** | -0.636 *** |
| Age | 0.069 *** | 0.054 *** | 0.066 *** | 0.047 ** |
| Age^2 | -0.002 *** | -0.001 *** | -0.001 *** | -0.001 ** |
| SD (Intercept) | 0.584 | 0.576 | 0.565 | 0.565 |
| SD Observation | 0.810 | 0.808 | 0.795 | 0.795 |
| Smoking |  | 0.014 *** | 0.014 *** | 0.025 *** |
| Age: Sexual Identity |  |  |  |  |
| Gay |  |  | -0.014 |  |
| Straight |  |  | -0.017 *** |  |
| Age: Smoke: Sexual Identity |  |  |  |  |
| Gay |  |  |  | -0.000 |
| Straight |  |  |  | -0.001 |
| Bisexual |  |  |  | 0.001** |
| **Random Effects** |  |  |  |  |
| $\sigma^2$ | 0.66 | 0.65 | 0.63 | 0.63 |
| $\tau_{00 \text{ id}}$ | 0.34 | 0.33 | 0.32 | 0.32 |
| ICC | 0.34 | 0.34 | 0.34 | 0.34 |
| $N_{\text{id}}$ | 2680 | 2680 | 2619 | 2619 |
| LogLik | -14477.651 | -14350.937 | -13190.159 | -13198.471 |
| AIC | 28965.302 | 28713.875 | 26396.318 | 26414.941 |
| BIC | 29001.701 | 28757.519 | 26453.923 | 26479.748 |
| Observations (N) | 10720 | 10657 | 9904 | 9904 |
| Marginal R<sup>2</sup> /Conditional R<sup>2</sup> | 0.002 / 0.343 | 0.010 / 0.343 | 0.014 / 0.345 | 0.014 / 0.345 |

Note: \*\*\* p < 0.001; \*\* p < 0.01; \* p < 0.05

## Discussion

The goal of our analyses was to determine the relationship between smoking behavior and depressive symptom trajectories across the life course, and how sexual identity moderates that relationship in a representative sample of U.S. adolescents. We determined the best fitting model was Model 2, which concludes that smoking behavior influences depression scores. Age, smoking behavior, and sexual identity all interactively impact depression scores across adolescence and adulthood. Integrating a person-centered approach in our analyses allowed us to focus on individuals and their own unique experiences and characteristics, which led to our findings of bisexual individuals having a significantly different impact of cigarette use on depressive symptoms across developmental age. Rather than presenting SM as one group, our results demonstrate the importance of separating sexual identities into their own respective groups (bisexual, lesbian, gay, etc.) as they may experience a significantly different effect. SM are often overlooked in research, and while this may be due to a lack of representation within a sample, that does not diminish the importance of their inclusion.

### Study Strengths

This is the first study to examine how cigarette use and behavior predict depression score trajectories across the lifespan and how sexual identification changes that relationship. This study fills a gap in the literature and provides findings that can be used to promote interventions to minimize disparities and promote health equity. A strength is the recognition of the heterogeneity of individuals and the utilization of a person-centered approach. Specifically, regarding depression, individuals likely have different patterns of symptoms that would not be captured by average effects alone. We also aimed to understand differences among subgroups of adolescents by sexual identity, which enables us to establish distinct targeted interventions for the specific needs of each group. By the study findings, for bisexual adolescents, the relationship between age and smoking status is positive and statistically significant, demonstrating a need for tailored interventions for this group to ensure improved depressive symptoms across the lifespan.

### Limitations & Directions for Future Research

As this is the first study of its kind, replication with a different cohort of young people would provide more evidence and validity of the presented results. Confounding is another limitation. It is important to consider outside variables that could be related to the study variables, such as socioeconomic status, other substance use/misuse, social support, history of trauma or abuse, etc. Additionally, our study did not consider the impact of intersectional factors (beyond age) that might be important. For example, SM women report greater disparities in cigarette smoking and poor mental health than SM men.^4,16,25^ Future studies with larger samples should consider interactions by sexual identity, gender, and race/ethnicity, for example. The Add Health sample also included too small a number of transgender and nonbinary young people to assess effects for gender minoritized (GM) groups; emerging research suggests that GM populations experience disparities in both smoking and poor mental health.^3,26^ Thus, including these groups in future research is critical for improving health equity.

## Conclusion

This study aimed to examine the relationship between smoking behavior and depression symptom trajectories across developmental age and how sexual identity moderates the relationship. The study findings suggest there is a statistically significant difference between age, cigarette use, and depressive symptoms in those who identify as bisexual. This leads to the conclusion that more early-life interventions are needed to support minoritized populations, specifically those identifying as bisexual. More research is imperative to better serve SM populations, and the implementation of specialized polices to improve mental health and smoking cessation for those who identify as bisexual is warranted.

The authors declare that they have no known competing financial interests or personal relationships that could have appeared to influence the work reported in this paper.

**Figure 1:**
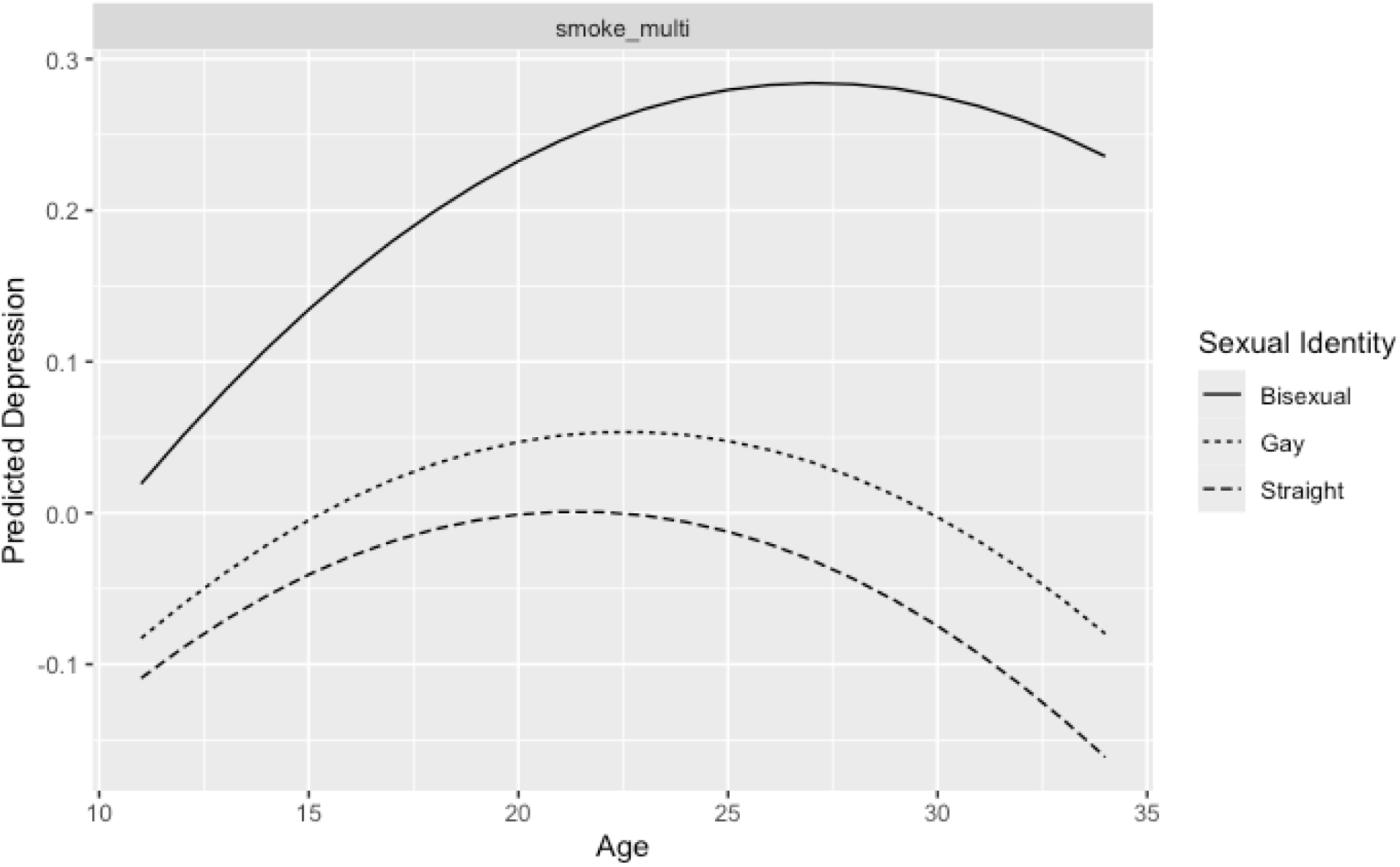
Predicted Depression Varying by Age and Smoking Level Grouped by Sexual Identity *Smoke_multi refers to the number of cigarettes smoked and frequency of smoking within the past 30 days

## Data Availability

All data produced in the present study are available upon reasonable request to the authors.

https://dataverse.unc.edu/dataverse/addhealth

